# Quality of Life and Associated Factors Among Stroke Survivors in Addis Ababa, Ethiopia: A Cross-Sectional Study

**DOI:** 10.64898/2026.01.21.26344563

**Authors:** Fitsum Zekarias, Fasika Molla, Dan Alemayehu, Edris Alemu, Getahun Gebre Bogale

## Abstract

**Introduction:** Stroke remains a leading cause of morbidity and mortality worldwide, with survivors often confronting significant physical, psychological, and social challenges. Quality of life (QoL) serves as a crucial indicator of recovery and long-term well-being among stroke survivors. However, little is known about the QoL of stroke survivors in Ethiopia. This study therefore aimed to assess QoL and associated factors among stroke survivors receiving care at specialized governmental hospitals in Addis Ababa, Ethiopia, in 2025.

**Methods:** An institution-based cross-sectional study was conducted from March 1 to May 1, 2025, among stroke survivors attending outpatient clinics and rehabilitation services at specialized governmental hospitals in Addis Ababa. A systematic random sampling technique was used to recruit 423 participants. Data were collected through interviewer-administered standardized questionnaires, coded, entered into EpiData version 4.7, and analyzed using SPSS version 21. Linear regression models were fitted, with unstandardized β coefficients and 95% confidence intervals (CI) reported. Statistical significance was set at a p-value < 0.05.

**Result:** The overall mean QoL score was 11.5 (SD = 2.89), with 44.6% of survivors scoring above the mean across all domains. Domain-specific mean scores were highest for upper extremity function (M = 16.8, SD = 4.99), followed by self-care (M = 14.9, SD = 4.64), social role (M = 14.8, SD = 4.89), and mobility (M = 14.6, SD = 6.97). The lowest score was recorded in the energy domain (M = 6.0, SD = 0.81). In the adjusted model, older age (β = –0.36, 95% CI: –0.51, –0.21), lower educational attainment (β = –0.34, 95% CI: –1.55, –0.26), rural residence (β = –0.57, 95% CI: –1.12, –0.33), depression (β = –0.55, 95% CI: –1.16, –0.35), and the presence of comorbidities (β = –0.67, 95% CI: –1.31, –0.11) were all significantly associated with lower QoL.

**Conclusion:** The overall quality of life of stroke survivors in Addis Ababa was poor. The findings underscore the multifactorial nature of post-stroke QoL and point out the need for integrated, patient-centered care. Interventions should prioritize early mental health screening, management of comorbidities, educational support, and improved access to rehabilitation services, particularly for elderly and rural populations, to enhance survivor well-being.

## Introduction

Stroke remains a major global health challenge and a leading cause of disability and death worldwide (1,2). It is clinically defined as an episode of acute neurological dysfunction caused by focal cerebral infarction or hemorrhage, with symptoms persisting for 24 hours or until death (3). Its suddenness often leaves individuals, families, and healthcare systems unprepared to manage its long-term sequel (4). Although advances in acute medical care have improved survival rates, stroke survivors frequently experience lasting physical, cognitive, and psychological impairments that profoundly diminish their health-related quality of life (QoL) (5,6).

Globally, an estimated 15 million people experience a stroke each year, with nearly 30% surviving with permanent disability (7). The economic burden of stroke is also substantial and projected to reach US$1 trillion by 2030 (2). According to the most recent Global Burden of Disease (GBD) estimates, the incidence of stroke has risen sharply, by approximately 70%, over the past two decades (8,9). Notably, the bulk of this burden resides in low– and middle-income countries (LMICs), which account for about 87.2% of stroke-related deaths and 89.4% of DALYs (10). While this disparity is profound, stroke also remains a major cause of death and disability in high-income countries as well. In the European Union, for instance, stroke remains the second leading cause of death and a primary cause of disability, affecting approximately 1.1 million individuals and resulting in 440,000 deaths annually (11). Ethiopia exemplifies the growing crisis in LMICs, reporting over 52,548 stroke incidents and 38,353 deaths in 2016, alongside hospital mortality rates ranging from 11% to 44% (12). This trend is mainly fueled by aging populations, the rising prevalence of risk factors such as hypertension and diabetes, smoking, and rapid urbanization (13).

Quality of life is a multidimensional concept, signifying an individual’s perception of their position in life within the context of their culture and value systems (5,14,15). Within this sphere, Health-Related Quality of Life (HRQoL) encompasses physical health, functional ability, mental state, and social well-being, serving as a crucial indicator of recovery among stroke survivors (5,6,15).

While a decline in Health-Related Quality of Life (HRQoL) following a stroke is a common phenomenon, its severity varies markedly between survivors in high-income countries (HICs) and low-and-middle-income countries (LMICs). While studies from HICs report significant declines, the measured impairment is generally less severe than in LMICs, as illustrated by an EQ-5D index of 0.757 in South Korea and a 5-year utility score of 0.50 in Australia (16,17). Conversely, more profound impairments were observed among stroke survivors in LMICs, particularly in Africa, evidenced by domain scores as low as 29% in Kenya, a global HRQoL score of 66.1 in Nigeria, and domain scores between 57.7% and 80.0% in Ghana (18–20). A regional systematic review further corroborates this pattern, finding typical mean scores between 5 and 12 (21). Within the specific context of Ethiopia, a 2021 study revealed that role limitations posed the greatest challenge to survivors, with the lowest mean scores observed for limitations due to physical health (15.69) and emotional problems (19.81) (22).

Quality of life following stroke is influenced by a complex interplay of sociodemographic, clinical, and psychosocial factors, including age, educational attainment, sex, residence (23–37), functional disability, comorbid conditions such as hypertension and diabetes (18,22,31,34,38,39), and depression (36,40–42).

In summary, stroke significantly impairs the quality of life of survivors, particularly in resource-limited settings like Ethiopia. A comprehensive understanding of the multifaceted factors, including age, educational status, rural residence, comorbidities, and depression, that influence QoL is therefore essential. Despite this, research into post-stroke HRQoL in sub-Saharan Africa is critically lacking. This was evident after conducting a comprehensive review of stroke literature conducted between 1970–2017, which showed that only 10 out of 51 studies addressed HRQoL, with just one conducted in Ethiopia.

Therefore, this study is significant as it aims to bridge the literature gap regarding HRQoL and associated factors among stroke survivors in specialized governmental hospitals in urban areas like Addis Ababa, Ethiopia. By identifying the most affected domains of QoL and the specific factors involved, the findings will provide critical insights for public health facilities and healthcare providers. This knowledge will also be crucial for the early identification of high-risk individuals and the development of targeted interventions to support recovery and improve overall well-being. Furthermore, the study’s findings are expected to inform program managers and policymakers in designing, implementing, and evaluating effective programs to enhance the physical, psychological, and social well-being of stroke survivors in Ethiopia.

## Methods and materials

### Study design, area, and period

This institution-based cross-sectional study was conducted in Addis Ababa, Ethiopia, between March 1 and May 1, 2025. Addis Ababa is a chartered city that has three layers of government: one municipality, ten sub-cities, and 116 districts/Woredas. The total area of the city is estimated at 526.99 km². The city serves as a social, economic, and political center of the country. As of August 2019, the total number of people living in the city is estimated at 4,592,000, with an annual growth rate of 4.4% (CSA 2019). The city has a total of 51 hospitals. Thirteen of these are public hospitals; six fall under the Addis Ababa Regional Health Bureau jurisdiction, and five are specialized referral hospitals. Among the specialized hospitals, three are administered at the federal level, while two are under the Addis Ababa Regional Bureau jurisdiction. Additionally, the city has 84 health centers and approximately 700 private clinics.

### Populations

All stroke survivors receiving care at any of the specialized government hospitals in Addis Ababa constituted the source population for this study. The study population comprised those stroke survivors who were selected through the sampling process.

### Sample size

The required sample size for assessing the QoL of stroke survivors receiving care at specialized government hospitals in Addis Ababa was determined using the single population proportion formula. The calculation assumed a proportion (p) of 0.5, anticipating that 50% of all stroke survivors receiving care at government specialized hospitals in Addis Ababa would score below the mean SS-QoL score, along with a 95% confidence interval and a 5% margin of error. This yielded an initial estimate of 384 participants. To account for potential non-response, this figure was increased by 10%, resulting in a final target sample size of 423. The sample size is also sufficient to assess factors significantly associated with QoL among stroke survivors, which include age, quality of rehabilitation service, and comorbidity (25,38).

### Sampling technique and procedures

In this study, a systematic random sampling procedure was followed to select the study participants. This involved proportionally allocating the sample size across the five specialized government hospitals available in Addis Ababa (Black Lion Hospital, Petrous Hospital, Minilik Hospital, and St. Paul Hospital, and Alert Hospital) based on their client load. Then, using client lists obtained from each hospital as sampling frames, the final participants were selected systematically (**Fig 1**).

**Fig 1.**
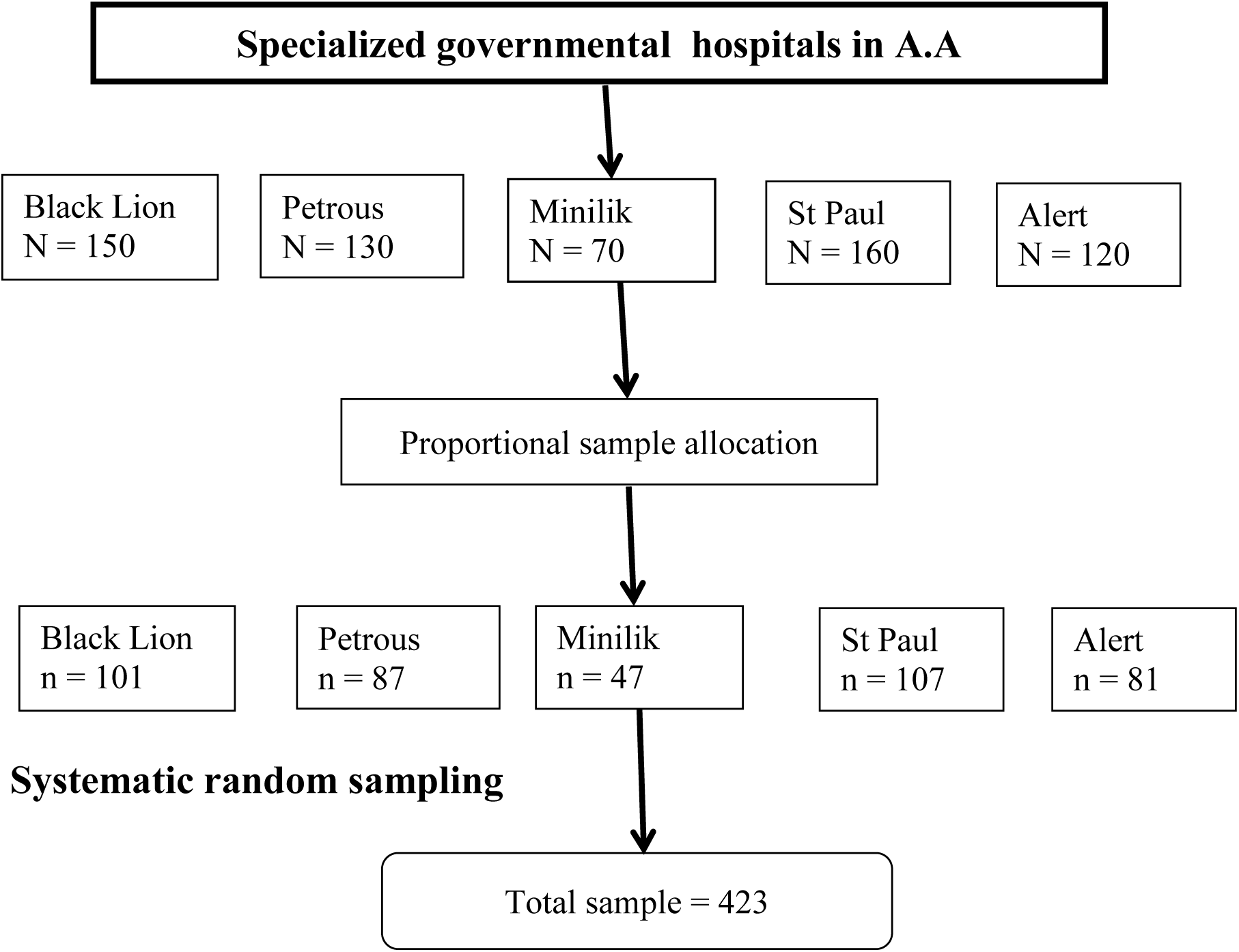
Sampling procedure followed to select stroke survivors in Addis Ababa, Ethiopia, 2025.

### Operational definitions

#### Quality of Life (QoL)

Quality of Life was measured using the Stroke-Specific Quality of Life scale (SS-QoL), a validated, self-reported outcome measure designed for stroke survivors. The 49-item questionnaire evaluates 12 domains of functioning and well-being. Each item is scored on a 5-point Likert scale. An overall SS-QoL score and individual domain scores were computed by summing the relevant items, with higher scores denoting better quality of life (43).

#### Comorbidity

Comorbidity was defined as the presence of one or more clinically diagnosed chronic conditions in addition to the index stroke, consistent with research linking multimorbidity to health outcomes (44).

#### Disability

Post-stroke functional status was assessed using the modified Rankin Scale (mRS), a standardized measure of global disability. The scale ranges from 0 (no symptoms) to 6 (death). For analytical purposes, a score of ≥2, indicating at least slight disability where a person is able to look after their own affairs without assistance but unable to carry out all previous activities, was used to define the presence of clinically significant disability (45).

#### Depression

Symptoms of depression were measured using the Depression subscale of the Hospital Anxiety and Depression Scale (HADS-D) (46). The total score was interpreted using established bands: a score of 0-7 indicates no depression, 8-10 indicates mild depression, 11-15 indicates moderate depression, and a score of 16 or higher suggests severe depression.

#### Social support

Perceived social support was measured using the Oslo Social Support Scale (OSSS-3) (47). The total score from its three items was categorized into three levels: a score of 3-8 indicates poor social support, 9-11 indicates moderate support, and 12-14 indicates strong social support.

### Data collection tool and technique

Data were collected via face-to-face interviews using a semi-structured questionnaire and a clinical data extraction checklist. The instrument comprised three main sections: socio-demographic variables (sex, age, marital status, religion, occupation, residence, and income); clinical variables (duration since stroke, type of stroke, affected site, comorbidity, disability, and rehabilitation service utilization); psychosocial variables (social support and depression); and QoL variables.

### Measurements and scoring

#### Stroke-Specific Quality of Life (SS-QoL)

This 49-item scale evaluates 12 domains: mobility, energy, upper extremity function, work/productivity, mood, self-care, social roles, family roles, vision, language, thinking, and personality. Items are rated on a 5-point Likert scale (1–5), with higher scores indicating better function or quality of life (43).

#### Hospital Anxiety and Depression Scale (HADS)

This 14-item tool screens for symptoms of anxiety and depression. It contains two 7-item subscales (HADS-A for anxiety and HADS-D for depression), which were analyzed separately (46).

#### Social Support

This was measured using the 3-item Oslo Social Support Scale (OSSS-3) (47).

### Quality assurance

The original English and validated Amharic versions were translated into Afan Oromo by linguistic and clinical experts. This version was then back-translated into English to ensure conceptual equivalence and accuracy before use in the study.

Prior to the main data collection, a pretest was conducted with 5% of the total calculated sample size at a hospital not included in the study (Zewditu Hospital). This pilot tested the questionnaire’s simplicity, flow, and consistency, leading to necessary refinements. For the main study, five trained nursing professionals administered the interviews, with supervision conducted by one public health officer and the principal investigator. All personnel, including the data collectors and supervisors, also took part in a two-day training session covering the study objectives, relevance, confidentiality protocols, participant rights, the informed consent process, and interview techniques. During the data collection, the principal investigator and supervisors also performed daily checks of all questionnaires for completeness. Finally, a comprehensive data cleaning and cross-checking procedure was conducted before performing the statistical analysis.

### Data processing and analysis

After the data were checked for completeness and rigorously cleaned, it were entered into EpiData version 4.7.0 before being exported to SPSS version 21 for analysis. Descriptive statistics (frequencies, percentages, means, and standard deviations) summarized the socio-demographic, clinical, and psychosocial characteristics of the sample and evaluated the distribution of responses.

Before conducting the linear regression analysis, the model’s key assumptions were validated. The assumptions of linearity and homoscedasticity were assessed by residual versus fitted value plots, which revealed no funnel shape. The normality of residuals was confirmed via a histogram and the Shapiro-Wilk test (p = 0.183). The Durbin-Watson statistic of 1.94 indicated no significant autocorrelation. Furthermore, all variables demonstrated Variance Inflation Factor (VIF) values below 4, confirming the absence of multicollinearity. Additionally, the internal consistency of the questionnaire was found to be acceptable, with a Cronbach’s alpha of 0.7.

### Ethical considerations

Ethical approval for this study was obtained from the Institutional Ethical Review Board (IRB) of the Asrat Woldeyes Health Science Campus, Debre Berhan University. Official permission letters were then secured from the administrative bodies of the selected hospitals. Written informed consent was obtained from all participants after explaining the study’s purpose. Participants were informed that their involvement was entirely voluntary, that they could withdraw at any time without consequence, and that all data would remain confidential. Privacy was maintained throughout the study, with only participant identification numbers used on data collection forms.

## Results

### Socio-demographic characteristics

A total of 416 stroke survivors participated in the study, yielding a response rate of 98.3%. The mean age of the survivors was 65 years (SD = 12.4). The majority were male (61.8%), married (56.7%), and lived in urban areas (89.4%). In terms of education and occupation, the largest groups held a diploma certificate (29.8%) and were unemployed (49.5%). Most survivors also identified as Orthodox Christian (59.4%). Regarding monthly income, majority earned between 1,000 and 5,000 Ethiopian Birr (39.4%) (**Table 1**).

**Table 1.**
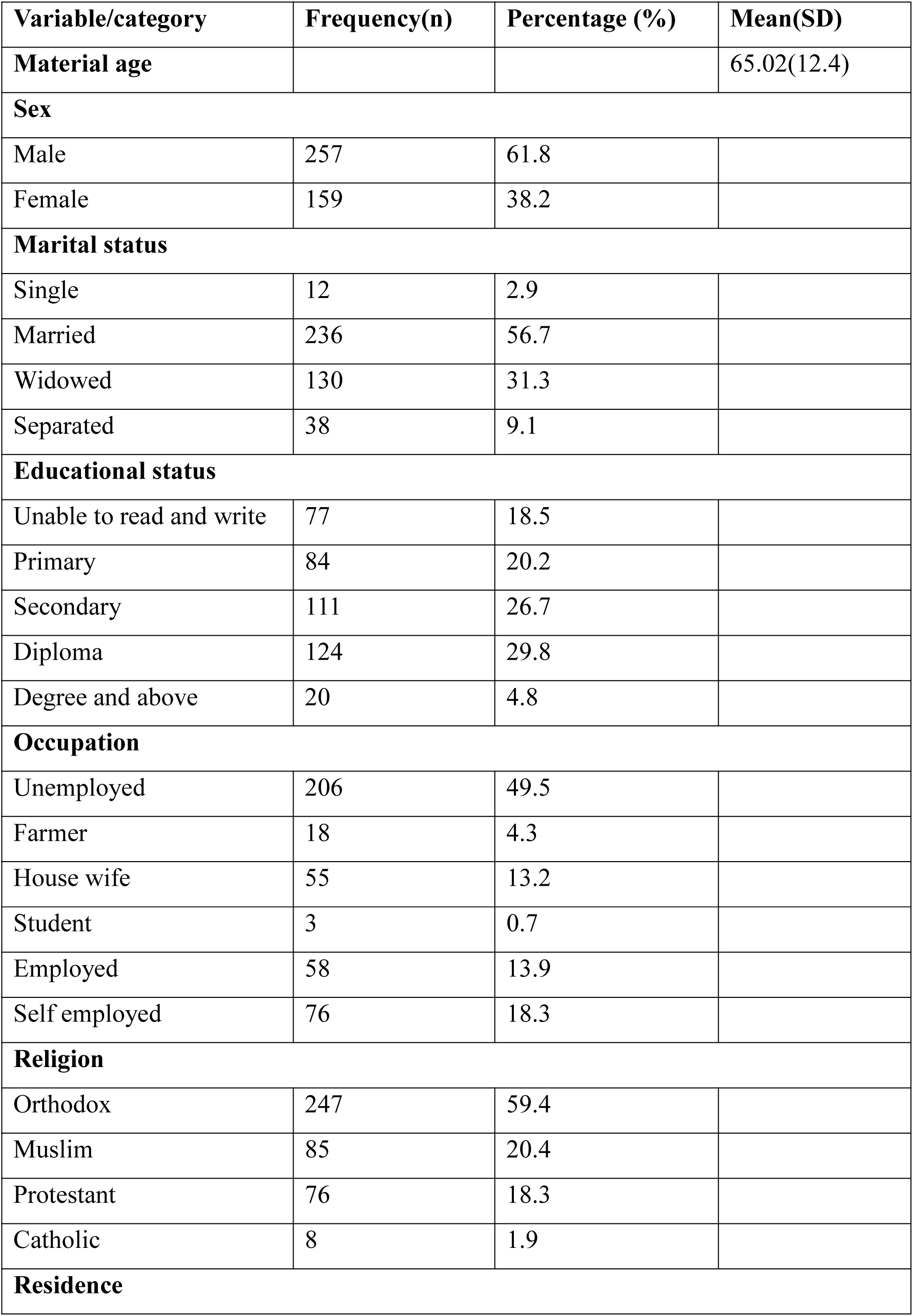
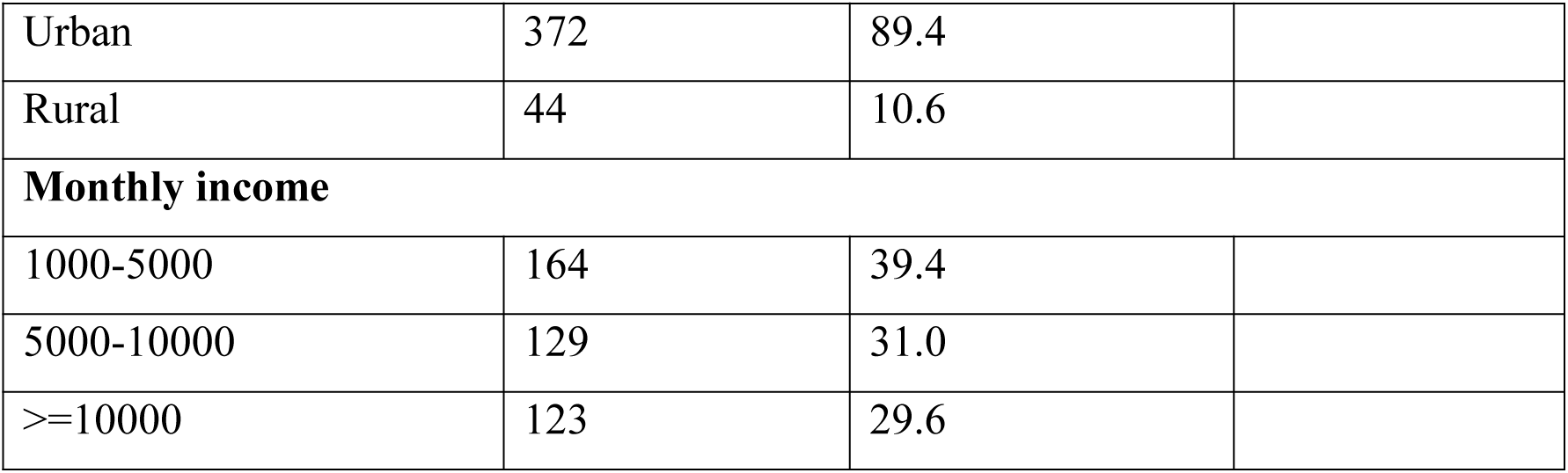
Socio-Demographic characteristic of stroke survivors in Addis Ababa, Ethiopia, 2025.

### Clinical factors

The mean time since stroke onset was 2.39 years (SD = 2.10). Comorbid conditions were prevalent, with 92.3% of survivors reporting at least one; among these, hypertension was the most common (47.9%). Ischemic stroke accounted for 75.5% of the cases, and the right side of the body was more frequently affected (79.8%) (**Table 2**).

**Table 2.** Clinical related factors among stroke survivors in Addis Ababa, Ethiopia, 2025.

| Variable/category | Frequency(n) | Percentage (%) | Mean(SD) |
| --- | --- | --- | --- |
| **Comorbidity** |  |  |  |
| Yes | 384 | 92.3 |  |
| No | 32 | 7.7 |  |
| **Comorbidity type** |  |  |  |
| HTN | 184 | 47.9 |  |
| DM | 106 | 27.6 |  |
| DM&HTN | 63 | 16.4 |  |
| Cardiac & HTN | 31 | 8.1 |  |
| **Type of stroke experienced** |  |  |  |
| Ischemic stroke | 314 | 75.5 |  |
| Hemorrhage stroke | 102 | 24.5 |  |
| **Side of the body affected** |  |  |  |
| Right | 332 | 79.8 |  |
| Left | 84 | 20.2 |  |
| **Duration of stroke in year** |  |  | 2.39 (2.10) |

### Psychosocial factors

Regarding the survivors’ mental state, 44.7% were not depressed, 32.7% were borderline depressed, and 22.6% were depressed. In terms of social support, 52.2% of survivors received poor support, 42.5% received intermediate support, and only 5.3% received strong support (**Table 3**).

**Table 3.** Psychosocial related factors among stroke survivors in Addis Ababa, Ethiopia, 2025.

| Variable/category | Frequency(n) | Percentage (%) |
| --- | --- | --- |
| **Depression** |  |  |
| Normal | 186 | 44.7 |
| Borderline | 136 | 32.7 |
| Depressed | 94 | 22.6 |
| **Social Support** |  |  |
| Poor | 217 | 52.2 |
| Intermediate | 177 | 42.5 |
| Strong | 22 | 5.3 |

### Quality of life

Stroke-specific quality of life was evaluated across various domains. In the energy domain, a significant proportion of the survivors reported experiencing high levels of fatigue, with 58.9% indicating they felt tired most of the time and 94.2% needing to stop and rest during the day. Within the family role domain, the majority reported that their physical condition adversely affected their personal lives (72.4%) and perceived themselves as a burden to their families (71.4%). Mild language impairments were also prevalent, with approximately half of the survivors (50%) experiencing slight difficulties during speaking and 40.6% reporting challenges in being understood. Substantial mobility impairments were observed, with over half unable to climb stairs (51.0%) or maintain balance while bending (50.5%). Regarding mood, 46.4% of the survivors reported a feeling of discouragement about the future and low self-confidence. In the personality domain, a majority indicated increased irritability (60.8%) and impatience (55.8%). In terms of self-care, most required considerable assistance with food preparation (51.2%), while a notable proportion needed some help with bathing (39.4%) and toileting (40.6%). Restrictions in social roles were evident, with 55.0% reporting that their physical condition interfered with their social life. Cognitive deficits were also apparent, as 58.9% experienced difficulties with memory. Mild impairments in upper extremity function, particularly fine motor tasks, were common. Vision problems were generally reported as mild, whereas work and productivity were frequently hindered, especially regarding daily household activities (41.6%) (**Sup file**). Overall, the mean QoL score was 11.5 (SD = 2.89), with 44.6% (95% CI: 40.1–49.8) of the survivors scoring above the mean across all domains (**Table 4**).

**Table 4.** Domain specific and overall mean quality of life scores of stroke survivors in Addis Ababa, Ethiopia, 2025.

| Domain | Mean (SD) | 95% CI |
| --- | --- | --- |
| Energy | 6.0 (0.81) | 5.8-6.2 |
| Family role | 7.1(2.23) | 6.1-7.4 |
| Language | 14.2(4.77) | 13.8-14.8 |
| Mobility | 14.6 (6.97) | 13.9-15.3 |
| Mood | 14.5(4.55) | 13.9-15.0 |
| Personality | 8.1(2.81) | 7.2-8.5 |
| Self-care | 14.9(4.64) | 13.5-15.4 |
| Social role | 14.8(4.89) | 13.8-15.3 |
| Thinking | 8.4(2.94) | 7.5-8.9 |
| Upper extremity function | 16.8(4.99) | 15.8-17.2 |
| Vision | 10.1(3.13) | 9.5-10.4 |
| Work productivity | 8.5(2.90) | 8.1-8.7 |
| Over mean score | 11.5(2.89) | 11.1-11.8 |
| Overall participant who scored above the mean | 44.6% | 40.1-49.8 |

### Factors associated with post-stroke quality of life

Variables that underwent simple linear regression analysis with a p-value score set at <0.2 include marital status, educational status, residence, occupation, depression, comorbidity, side of body affected, and stroke type. The model explained 68.6% of the variance, and all variance inflation factors were below 4. Model fit was validated (F = 11.46, p = 0.01). A subsequent multiple linear regression model with backward elimination identified factors significantly associated with quality of life (p < 0.05): age, educational status, residence, depression, and comorbidity.

In detail, for every one-year increase in age, the HRQoL score decreased by 0.36 units (β = –0.36, 95% CI: –0.51 to –0.21, p = 0.02), holding other variables constant. Stroke survivors who were unable to read or write had an HRQoL score 0.34 units lower than those with a degree or higher education (β = –0.34, 95% CI: –1.55 to –0.26, p = 0.03). Participants residing in rural areas also scored 0.57 units lower than those in urban areas (β = –0.57, 95% CI: –1.12 to –0.33, p = 0.021). Those who were depressed scored 0.55 units lower than those who were not (β = –0.55, 95% CI: –1.16 to –0.35, p = 0.003). Finally, stroke survivors with comorbid conditions scored 0.67 units lower than those without comorbidities (β = –0.67, 95% CI: –1.31 to –0.11, p = 0.011), with other variables held constant (**Table 5**).

**Table 5.** Multiple linear regression associated with stroke survivor quality of life in Addis Ababa, Ethiopia, 2025.

| Variable | Category | $\beta$ coefficient | 95% CI | P-value |
| --- | --- | --- | --- | --- |
| Age in year |  | -0.36 | (-0.51, -0.21) | 0.02* |
| Marital status | Single | -6.66 | (-8.5, 1.16) | 0.44 |
|  | Separated | -4.67 | (-6.19, 2.06) | 0.79 |
|  | Widowed | 0.62 | (-10.99, 1.24) | 0.32 |
|  | Married | 1 |  |  |
| Educational status | Unable to read and write | -0.34 | (-1.55, -0.26) | 0.03* |
|  | Primary | 0.47 | (-0.89, 3.37) | 0.55 |
|  | Secondary | -3.86 | (-8, 11, 4, 95) | 0.76 |
|  | Diploma | -4.55 | (-10.63, 3.80) | 0.24 |
|  | Degree and above | 1 |  |  |
| Occupation | Unemployed | 0.34 | (-2.86 to 3.53) | 0.84 |
|  | Farmer | -1.69 | (-6.10 to 2.72) | 0.45 |
|  | House wife | 1.05 | (-9.42, 2.77) | 0.66 |
|  | Student | -5.44 | (-8.53,1.78) | 0.93 |
|  | Self employed | 1.46 | (-0.93,2.33) | 0.12 |
|  | Govt employed | 1 |  |  |
| Residence | Rural | -0.57 | (-1.12, -0.33) | 0.021* |
|  | Urban | 1 |  |  |
| Depression | Depressed | -0.55 | (-1.16, -0.35) | 0.003** |
|  | Borderline depressed | 1.06 | (-0.85,1.30) | 0.676 |
|  | Not depressed | 1 |  |  |
| Comorbidity | Yes | -0.67 | (-1.31, -0.11) | 0.011* |
|  | No | 1 |  |  |
| Side of the body affected | Right | -1.49 | (-2.37, 0.62) | 0.56 |
|  | Left | 1 |  |  |
| Types of the stroke | Ischemic | 0.87 | (-0.36- 1.12 ) | 0.79 |
|  | Hemorrhage | 1 |  |  |

## Discussion

This study assessed quality of life (QoL) and associated factors among stroke survivors attending specialized hospitals in Addis Ababa, Ethiopia. The overall mean QoL score was 11.5 (SD = 2.89), with 44.6% (95% CI: 40.1–49.8) of the survivors scoring above the mean across all domains.

This finding aligns with studies from similar settings. For instance, research in southeast Nigeria reported lower mean HRQoL scores (ranging from 7 to 16) among stroke survivors compared to healthy controls (31). Furthermore, a systematic review and meta-analysis of 28 studies across eight African countries concluded that stroke survivors consistently report low HRQoL (mean scores between 5 and 12), with significant deficits compared to healthy populations (21). This similarity may be attributed to shared challenges in low-resource settings, including limited access to comprehensive rehabilitation, financial constraints, inadequate post-discharge support, and cultural stigma surrounding disability, all of which can hinder the resumption of daily activities and social roles (31).

The quality of life of stroke survivors in Addis Ababa was better than those in Sierra Leone, which demonstrated a mean quality-adjusted life year (QALY) score of 0.28 (SD = 0.35) one year post-stroke. The authors attributed this low QALY to higher stroke severity, delayed hospital presentation, and a lack of specialized stroke services (35). Conversely, the quality of life of stroke survivors in Addis Ababa was worse than those from higher-resource settings. The stroke survivors in South Korea, for instance, demonstrated better QoL scores compared to the stroke survivor in Addis Ababa, which was reflected in their EQ-5D index score of 0.757 ± 0.012 (17). Similarly, stroke survivors in Ghana demonstrated higher domain-specific scores, with cognitive function at 57.7% and spiritual well-being at 80.0%, indicating that while some domains may be preserved, cognitive impairment is a common post-stroke phenomenon (18). These disparities likely stem from better-developed rehabilitation services, comprehensive health insurance, and established community-based care systems that facilitate early intervention and long-term follow-up, resources often scarce in low-income countries (48). The stroke survivors in Kenya also demonstrated better QoL scores across physical (29.9%) and psychological (48.5%) domains compared to our cohort (20). This difference may be explained by the younger age of the Kenyan participants, who potentially benefit from greater neuroplasticity, fewer comorbidities, and a higher capacity for physical and psychosocial adaptation following a stroke.

Our study also identified several factors significantly associated with HRQoL among stroke survivors. Among those, age was found to negatively correlate with HRQoL. This finding resonates with the findings from Tunisia (34), Sierra Leone (35), Sub-Saharan Africa (33), and Nigeria (31). This might be due to the fact that older adults have reduced physiological resilience, including weaker muscle strength and diminished neuroplasticity, hindering recovery. A higher prevalence of comorbidities like hypertension and diabetes in older age can further impede rehabilitation and lower QoL (34). Additionally, older survivors often face greater barriers to accessing rehabilitation services due to physical immobility, financial limitations, and inadequate caregiver support, leading to social isolation, depression, and ultimately poor QoL (33).

Educational status, specifically the inability to read and write, was another factor that negatively affected HRQoL. This finding aligns with the findings reported from Nigeria (32), Ethiopia (49), Kenya (50), Tunisia (34), and a systematic review of Sub-Saharan Africa (33). This might be explained by the fact that lower educational attainment limits health literacy, impairing an individual’s ability to understand medical advice, adhere to therapy, and the capacity to care for oneself, all crucial for stroke recovery. Furthermore, lower education is frequently linked to socioeconomic disadvantage, restricting access to healthcare services, assistive devices, and necessary support systems (31,50).

Our study revealed that rural residence lead to poorer HRQoL, which was also true according to the studies in Sierra Leone (35), Tanzania (36), and South Africa (37). Qualitative research conducted in rural Uganda also highlights the compounded challenges faced by stroke survivors, including limited access to rehabilitation, economic hardship, and insufficient social support (51). The disproportionate burden of stroke in rural South Africa has also been documented, exacerbated by lifestyle risk factors and scarce healthcare resources (52). These challenges, encompassing transportation barriers, financial strain, lower health literacy, and social isolation, can delay treatment, reduce rehabilitation adherence, and increase the risk of depression and disability (36,37,51).

Depression was another factor that was strongly associated with reduced HRQoL, a finding corroborated by studies in Ethiopia (41), Nigeria (42), China (40), Tunisia (34), and Sweden (39). This can be explained by the fact that depression can severely impact motivation, cognitive function, and adherence to rehabilitation protocols. Feelings of hopelessness and social withdrawal commonly experienced by depressed individuals can lead to poorer physical and emotional functioning, creating a detrimental cycle that impedes overall recovery and well-being (39).

Finally, the presence of comorbid conditions was linked to lower HRQoL. Studies in Ethiopia (22) and Ghana (18) have similarly shown that hypertension and diabetes are associated with worse outcomes across physical and cognitive domains. Research from Sweden also confirmed that a high comorbidity burden significantly increases the risk of poor long-term functional outcomes and mortality post-stroke (39). This might be due to the fact that comorbidities complicate clinical management, can increase initial stroke severity, prolong recovery, and elevate the risk of recurrent vascular events, thereby substantially diminishing quality of life (18).

## Strength and limitation of the study

The strengths of this study include the use of validated, standardized tools to assess quality of life, depression, and social support, ensuring reliable and comparable findings. However, its cross-sectional design precludes establishing causal relationships and tracking changes in quality of life over time. Furthermore, reliance on quantitative data limited the exploration of survivors’ subjective experiences and contextual factors. Therefore, future studies should employ longitudinal designs and qualitative methods, such as in-depth interviews, to address these gaps.

## Conclusion and recommendation

The overall quality of life (QoL) of stroke survivors who are receiving care at the specialized hospitals of Addis Ababa was poor. Key factors significantly associated with lower QoL were older age, lower educational attainment, rural residence, comorbid conditions, and depression. These findings underscore the multifaceted nature of post-stroke recovery and highlight the critical need for integrated, person-centered care strategies. To enhance the well-being of stroke survivors, we recommend the following coordinated actions. For stroke survivors, it is essential to actively participate in rehabilitation and follow-up care to enhance recovery, diligently manage comorbid conditions like hypertension and diabetes to prevent health deterioration, and seek timely psychological support for depressive symptoms to safeguard mental well-being and overall quality of life.

For healthcare professionals, we recommend integrating routine screening for depression and comorbidities into standard post-stroke follow-up protocols. Providing clear, culturally appropriate health education is crucial for survivors with limited literacy to promote self-care and treatment adherence. Furthermore, clinical attention and support should be prioritized for elderly survivors and those residing in rural areas, as these groups are at a significantly higher risk for poor long-term outcomes.

For hospitals and the broader health system, we recommend establishing comprehensive, multidisciplinary rehabilitation programs that include physiotherapy, occupational therapy, mental health services, and nutritional counseling. Strengthening referral linkages between tertiary city hospitals and rural primary healthcare units is necessary to ensure seamless continuity of care. Finally, we recommend implementing regular, evidence-based training for all relevant healthcare staff to ensure proficiency in contemporary stroke management and rehabilitation practices.

## Declarations

### Data sharing statement

The datasets used and/or analyzed during the current study are available from the corresponding author upon reasonable request.

### Consent for publication

Not applicable

## Data Availability

All data produced in the present study are available upon reasonable request to the authors

