## Supplementary material for "Quality of Life and Associated Factors Among Stroke Survivors in Addis Ababa, Ethiopia: A Cross-Sectional Study": Sup file

**Supplementary table**

| **Energy** | **Frequency (percentage)** | | | | | | | | | | | | | | |
| --- | --- | --- | --- | --- | --- | --- | --- | --- | --- | --- | --- | --- | --- | --- | --- |
| Item | Strongly Agree | | | Agree | | | | Neutral | Disagree | | | Strongly Disagree | | | |
| I felt tired most of the time. | 71(17.1) | | | 245(58.9) | | | | 24(5.75) | 45(10.8) | | | 31(7.45) | | | |
| I had to stop and rest during the day | 19(4.6) | | | 392(94.2) | | | | 2(0.5) | 2(0.5) | | | 1(0.2) | | | |
| I was too tired to do what I wanted to do | 57(13.7) | | | 307(73.8) | | | | 7(1.7) | 44 (10.6) | | | 1(0.2) | | | |
| **Family role** |  | | | | | | | | | | | | | | |
| Item | Strongly Agree | Agree | | | | Neutral | | | | Disagree | | | Strongly Disagree | | |
| I didn't join in activities just for fun with my family. | 22(5.3) | 241(57.9) | | | | 68(16.3) | | | | 82(19.7) | | | 3(0.7) | | |
| I felt I was a burden to my family | 30(7.2) | 297(71.4) | | | | 14(3.4) | | | | 72(17.3) | | | 3(0.7) | | |
| My physical condition interfered with my personal life. | 32(7.7) | 301(72.4) | | | | 12(2.9) | | | | 68(16.3) | | | 3(0.7) | | |
| **Language** |  | | | | | | | | | | | | | | |
| Item | No trouble at all | A little trouble | | | Some trouble | | | | | A lot of trouble | | | Couldn't do it at all | | |
| Did you have trouble speaking? For example, get stuck, stutter, stammer, or slur your words? | 23 (5.5) | 208(50) | | | 74(17.8) | | | | | 98(23.6) | | | 13(3.1) | | |
| Did you have trouble speaking clearly enough to use the telephone? | 17(4.1) | 196(47.1) | | | 70(16.8) | | | | | 116(27.9) | | | 17(4.1) | | |
| Did other people have trouble in understanding what you said? | 20(8.4) | 169(40.6) | | | 94(22.6) | | | | | 112(26.9 | | | 21(5) | | |
| Did you have trouble finding the word you wanted to say? | 14(3.4) | 157(37.7) | | | 94(22.6) | | | | | 124(29.8) | | | 27(6.5) | | |
| Did you have to repeat yourself so others could understand you? | 22(5.3) | 155(37.3) | | | 97(23.3) | | | | | 119(28.6) | | | 23(5.5) | | |
| **Mobility** |  | | | | | | | | | | | | | | |
| Item | No trouble at all | A little trouble | | | | | Some trouble | | | A lot of trouble | | | | Couldn't do it at all | |
| Did you have trouble walking? (If patient can't walk, go to question 4 and score questions 2-3 | 17(4.1) | 83(20.0) | | | | | 109(26.2) | | | 157(37.7) | | | | 50(12.0) | |
| Did you lose your balance when bending over to or reaching for something? | 14(3.4) | 55(13.2) | | | | | 123(29.6) | | | 14(3.4) | | | | 210 (50.5) | |
| Did you have trouble climbing stairs? | 9(2.2) | 63(15.1) | | | | | 124(29.8) | | | 8(1.9) | | | | 212(51) | |
| Did you have to stop and rest more than you would like when walking or using a wheelchair? | 12(2.9) | 116(27.9) | | | | | 94(22.6) | | | 159(38.2) | | | | 35(8.4) | |
| Did you have trouble with standing? | 24 (5.8) | 120(28.8) | | | | | 95(22.8) | | | 150 (36.1) | | | | 27 (6.5) | |
| Did you have trouble getting out of a chair? | 22(5.3) | 126(30.3) | | | | | 95(22.8) | | | 148(35.6) | | | | 25(6.0) | |
| **Mood** |  | | | | | | | | | | | | | | |
| Item | Strongly Agree | Agree | | | | Neutral | | | | Disagree | | | Strongly Disagree | | |
| I was discouraged about my future. | 20 (4.8) | 193(46.4) | | | | 59(14.2) | | | | 137(32.9) | | | 7(1.7) | | |
| I wasn't interested in other people or activities. | 16(3.8) | 176(42.3) | | | | 59(14.2) | | | | 160(38.5) | | | 5(1.2) | | |
| I felt withdrawn from other people. | 17(4.1) | 179(43.0) | | | | 44(0.6) | | | | 168 (40.4) | | | 8(1.9) | | |
| I had little confidence in myself. | 20(4.8) | 193(46.4) | | | | 33(7.9) | | | | 155(37.3) | | | 15(3.6) | | |
| I was not interested in food. | 13(3.1) | 178(42.8) | | | | 32(7.7) | | | | 181(43.5) | | | 12(2.9) | | |
| **Personality** |  | | | | | | | | | | | | | | |
| Item | Strongly Agree | Agree | | | | Neutral | | | | | Disagree | | Strongly Disagree | | |
| I was irritable. | 25 (6.0) | 253(60.8) | | | | 20(4.8) | | | | | 103(24.8) | | 15(3.6) | | |
| I was inpatient with others. | 17(4.1) | 232(55.8) | | | | 25(6.0) | | | | | 130(31.3) | | 12(2.9) | | |
| My personality has changed. | 18(4.3) | 218(52.4) | | | | 21(5.0) | | | | | 140(33.7) | | 19(4.6) | | |
| **Social role** |  | | | | | | | | | | | | | | |
| Item | Strongly Agree | Agree | | | | Neutral | | | | | Disagree | | Strongly Disagree | | |
| I didn't go out as often as I would like. | 25(8.0) | 156(37.5) | | | | 41(9.9) | | | | | 176(42.3) | | 18(4.3) | | |
| I did my hobbies and recreation for shorter periods of time than I would like. | 9(2.2) | 185(44.5) | | | | 38(9.1) | | | | | 157(37.7) | | 27(6.5) | | |
| I didn't see as many of my friends as I would like. | 11(2.6) | 187(45.0) | | | | 30(7.2) | | | | | 167(40.1) | | 21(5.0) | | |
| I had sex less often than I would like | 14(3.4) | 150(36.1) | | | | 82(19.7) | | | | | 141(33.9) | | 29(7.0) | | |
| My physical condition interfered with my social life. | 20(4.8) | 229(55.0) | | | | 18(4.3) | | | | | 124(29.8) | | 25(6.0) | | |
| **Self-care** |  | | | | | | | | | | | | | | |
| Item | Total help needed | | A lot of help | | Some help | | | | | A little help | | | | | No help needed |
| Did you need help preparing food? | 71(17.1) | | 213(51.2) | | 51(12.3) | | | | | 70(16.8) | | | | | 11(2.6) |
| Did you need help eating? For example, cutting food or preparing food? | 13(3.1) | | 141(33.9) | | 73(17.5) | | | | | 155(37.3) | | | | | 34(8.2) |
| Did you need help getting dressed? For example, putting on socks or shoes, buttoning buttons, or zipping? | 22(5.3) | | 126(30.3) | | 78(18.8) | | | | | 161(38.7) | | | | | 29(7.0) |
| Did you need help taking a bath or a shower? | 27(6.5) | | 113(27.2) | | 79(19.0) | | | | | 164(39.4) | | | | | 33(7.9) |
| Did you need help to use the toilet? | 22(5.3) | | 112(26.9) | | 83(20.0) | | | | | 169(40.6) | | | | | 30(7.2) |
| **Thinking** |  | | | | | | | | | | | | | | |
| Item | Strongly Agree | | Agree | | Neutral | | | | | Disagree | | | | | Strongly Disagree |
| It was hard for me to concentrate | 16(3.8) | | 222(53.4) | | 31(7.5) | | | | | 124(29.8) | | | | | 23(5.5) |
| I had trouble remembering things | 10(2.4) | | 245(58.9) | | 21(5.0) | | | | | 115(27.6) | | | | | 25(6.0) |
| I had to write things down to remember them. | 9(2.2) | | 228(54.8) | | 20(4.8) | | | | | 130(31.3) | | | | | 29(7.0) |
| **Upper extreme function** |  | | | | | | | | | | | | | | |
| Item | Couldn't do it at all | | A lot of trouble | | Some trouble | | | | | A little trouble | | | | | No trouble at all |
| Did you have trouble writing or typing? | 16(3.8) | | 177(42.5) | | 40(9.6) | | | | | 141(33.9) | | | | | 42(10.1) |
| Did you have trouble putting on socks? | 9(2.2) | | 113(27.2) | | 55(13.2) | | | | | 190(45.7) | | | | | 49(11.7) |
| Did you have trouble buttoning buttons? | 9(2.2) | | 110(6.4) | | 48(11.5) | | | | | 201(48.3) | | | | | 48(11.5) |
| Did you have trouble zipping a zipper? | 7(1.7) | | 100(24.0) | | 55(13.2) | | | | | 202(48.6) | | | | | 52(12.5) |
| Did you have trouble opening a jar? | 8(1.9) | | 92(22.1) | | 57(13.7) | | | | | 206(49.5) | | | | | 53(12.7) |
| **Vision** |  | | | | | | | | | | | | | | |
| Item | Couldn't do it at all | | A lot of trouble | | Some trouble | | | | | A little trouble | | | | | No trouble at all |
| Did you have trouble seeing the television well enough to enjoy a show? | 8(1.9) | | 111(26.7) | | 63(15.1) | | | | | 170(40.9) | | | | | 64(15.4) |
| Did you have trouble reaching things because of poor eyesight? | 8(1.9) | | 124(29.8) | | 70(16.8) | | | | | 153(36.8) | | | | | 61(14.7) |
| Did you have trouble seeing things off to one side? | 6(1.4) | | 123(29.6) | | 73(17.5) | | | | | 152(36.5) | | | | | 62(14.9) |
| **Work productivity** |  | | | | | | | | | | | | | | |
| Item | Couldn't do it at all | | A lot of trouble | | Some trouble | | | | | A little trouble | | | | | No trouble at all |
| Did you have trouble doing daily work around the house? | 14(3.4) | | 173(41.6) | | 108(26.0) | | | | | 102(24.5) | | | | | 19(4.6) |
| Did you have trouble finishing jobs that you started? | 27(6.5) | | 161(38.7) | | 112(27.4) | | | | | 95(22.8) | | | | | 19(4.6) |
| Did you have trouble doing the work you used to do? | 26(6.3) | | 155(37.3) | | 110(26.4) | | | | | 104(25.0) | | | | | 21(5.0) |
